# Heterogeneous but Segmentable: A Data-Driven Approach to Modelling Long-Term Care Trajectories in Multiple Sclerosis

**DOI:** 10.64898/2026.01.28.26345045

**Authors:** Märt Vesinurm, Laura Mäkitie, Paul Lillrank, Lauri Saarinen, Paulus Torkki, Sini M Laakso, Miika Koskinen

## Abstract

Managing chronic diseases with unpredictable care demand creates significant operational challenges for healthcare systems. Mapping long-term care trajectories is crucial for improving resource allocation, anticipating service needs, and designing efficient care pathways. We used a data-driven approach to map six-year care trajectories for 962 newly diagnosed multiple sclerosis patients, identify utilization clusters, and determine predictors of high utilization. We analyzed Event logs of remote, outpatient, emergency, and inpatient contacts from one year pre- to five years post-diagnosis using K-means clustering to identify utilization clusters, logistic regression to identify predictors of high utilization, and process mining to model variation between care trajectories. We identified two distinctive utilization clusters: a high-utilization cluster (14.1 % of patients) with persistently elevated annual encounter volumes across all care settings and low-utilization cluster (85.9 % of patients) with lower and declining use. Median service costs were €18,736 vs. €6,052 in high- and low-utilization clusters, respectively. Two or more early relapses were the strongest predictor of high utilization (OR = 6.33, 95 % CI 3.49-11.50, p < 0.001), with number of planned early remote and outpatient care contacts being also associated with future service utilization (OR = 1.07, 95 % CI 1.04–1.10, p < 0.001). High-utilization trajectories were approximately three times longer (82.4 vs 25.9 events) and more variable (3.1 vs 2.4 unique events per patient). These utilization clusters and their distinct trajectories provide a pragmatic segmentation of multiple sclerosis patients to support early identification of high-utilization subgroups and more robust capacity planning in specialist care.

**Highlights:**

- We tracked the care trajectories of 962 people with relapsing–remitting multiple sclerosis using a Finnish population-based specialist-care datalake covering both inpatient and outpatient neurology services.
- Patients fell into two distinct utilization clusters: a high-utilization cluster with frequent contacts across all care settings and a low-utilization cluster with lower and declining use.
- Two or more early relapses, and the number of early outpatient and remote contacts were strong predictors of a patient’s long-term affiliation in the high-utilization cluster.
- Segmented care trajectories showed that high-utilization patients followed longer, more varied, and acute-oriented care patterns and had much higher service encounter costs.
- These findings can help clinicians and managers identify potential high-utilization patients early, target resources more effectively, and plan for future healthcare demand.

## Introduction

Managing chronic diseases with unpredictable care demand poses persistent operational challenges for healthcare systems [1,2]. Patients require long-term, coordinated outpatient follow-up punctuated by unpredictable acute episodes that trigger urgent or inpatient care, generating volatile and difficult-to-forecast demand. This volatility undermines capacity planning, resource allocation, and the design of robust care pathways. From an operations perspective, distinguishing patients with persistently high service intensity from those with declining use is critical for proactive capacity planning and care pathway design.

Multiple sclerosis (MS) is a chronic, immune-mediated disorder of the central nervous system [3]. Finland is a high-risk region for MS [4] with a prevalence of 241.5 per 100,000 (overall global prevalence 35.9 per 100,000) and incidence of 8.6 per 100,000 (overall global incidence 2.1 per 100,000) [3,5]. MS is Finland’s third-leading reason for disability pensions, and its societal burden is substantial with an average annual cost per patient at approximately €50,000 (US $54,000) [6]. Because disability progression can be slowed but not reversed, people with MS need lifelong, multidisciplinary care. The primary therapeutic objective in MS is to prevent relapses, as each relapse not only accelerates long-term disability but is also a major driver of costly healthcare resource use. Consequently, MS represents a paradigmatic case for operational analysis: newly diagnosed patients show elevated emergency department (ED) use and inpatient episodes, yet their utilization patterns remain highly heterogeneous (3). Disease severity, comorbidity burden, and treatment adherence are key drivers of this heterogeneity and associated costs (4,5).

Using routine data collected in electronic health records (EHR), process mining (PM) has emerged as a data-driven approach for reconstructing and analyzing sequences of time-stamped events to reveal common pathways, deviations, and bottlenecks [7–9]. PM enables the discovery, monitoring, and improvement of actual care trajectories as documented in EHRs, rather than relying solely on predefined clinical pathways that may not reflect real-world practice [10,11]. Systematic reviews indicate that PM is increasingly used in healthcare for patient-flow discovery, guideline-conformance checking, and bottleneck analysis, often integrating domain expertise and operational performance indicators [7,10]. While most PM work in healthcare focuses on short-term processes such as patient flows within an ED or surgeries, applications in chronic conditions including oncology, cardiology, and mental health have demonstrated that PM and related sequence-analysis methods can map multi-year care trajectories and segment patients into distinct utilization patterns [12–18]. However, actionable managerial levers, the key deviation points, and the drivers of patient-level variation are largely unknown, which directly hinders efforts to design efficient care pathways, forecast demand, and allocate resources for chronic, unpredictable conditions such as MS. This study addresses that gap.

Building on emerging literature, prior PM and sequence-based studies in the context of MS have segmented patients into three to five clusters capturing gradations of care intensity or treatment patterns [19–22]. While these granular classifications highlight the heterogeneity of MS, their operational utility for health service management is constrained. The boundaries between, for example, medium and high-utilization trajectories often lack clear managerial meaning when translated into actionable resource allocation or pathway design decisions. From a planning perspective, overly detailed segmentation can obscure rather than clarify the key contrast that matters most for operational decision-making: the difference between patients whose care trajectories remain relatively stable and predictable, and those who generate persistently high or volatile service demand.

Here we use a data-driven approach to map the heterogeneity of care trajectories and develop a managerially useful segmentation of RRMS patients by applying PM to a six-year longitudinal cohort treated at a specialist care center that provides both in-hospital and outpatient services in a population-based setting. We (i) identify clusters of patients with similar care trajectories in terms of healthcare resource use; (ii) identify individual-level predictors of high resource use; and (iii) describe trajectories from one year pre-diagnosis to five years post-diagnosis for these clusters. These trajectory archetypes and their predictors offer actionable input for capacity planning, service design, and targeted interventions in MS care.

## Methods

This was a retrospective observational cohort study using electronic health record (EHR) data from the datalake of Helsinki University Hospital (HUS) covering the years 2008-2023. The cohort included all patients diagnosed with relapsing–remitting multiple sclerosis (RRMS) between 1.1.2009-31.12.2018. The study follows STROBE guidelines for observational research [23]. All analyses were performed in R 4.5.0 with RStudio 2024.12.1 (Posit Software, PBC).

### Empirical Context

Finland provides an interesting context for this study due to its distinctive healthcare system, which combines publicly funded and private services. Like in many other societies, the financing of healthcare presents ongoing challenges, yet Finland offers exceptionally comprehensive data on care processes and their effectiveness. This combination makes it a particularly suitable setting for examining the dynamics and outcomes of healthcare provision. In Finland, the diagnosis of MS is carried out exclusively within the public healthcare system without insurance-based access differences. In the Helsinki region, neurological care is centralized at HUS serving a population of 1.7 million. The diagnostic process typically includes a neurological exam, brain and spinal magnetic resonance imaging, and cerebrospinal fluid analysis [24].

Ongoing management of MS is structured around scheduled in-person and telephone consultations with neurologists and specialist nurses [25]. Most patients begin disease-modifying therapy shortly after diagnosis, which requires laboratory assessments both before and during treatment. A follow-up MRI is generally performed six months after treatment initiation, usually accompanied by a physician visit. Nurse appointments provide an opportunity to discuss practical aspects of medication use and monitoring. Newly diagnosed patients are also invited to attend twice-yearly group education sessions where healthcare professionals provide information on MS and patients can interact with peers. Over the disease course, routine neurologist visits are typically scheduled every 6–12 months, with additional appointments arranged as needed aligned with international treatment guidelines [26]. Patients can also contact their nurse between visits if new symptoms arise or further guidance is required. Finnish MS care broadly follows international practices. Distinctive features, such as the role of occupational health services and regional variation in rehabilitation access, may shape utilization patterns and should be considered when comparing across systems.

### Study Design and Setting

The study design followed a three-step workflow (Figure 1). We clustered patients based on their long-term healthcare utilization (A). Logistic regression was then used to predict cluster membership based on baseline characteristics (B). Finally, process mining (PM) was used to map care trajectories for the study period for the full cohort and stratified by cluster (C).

**Figure 1.**
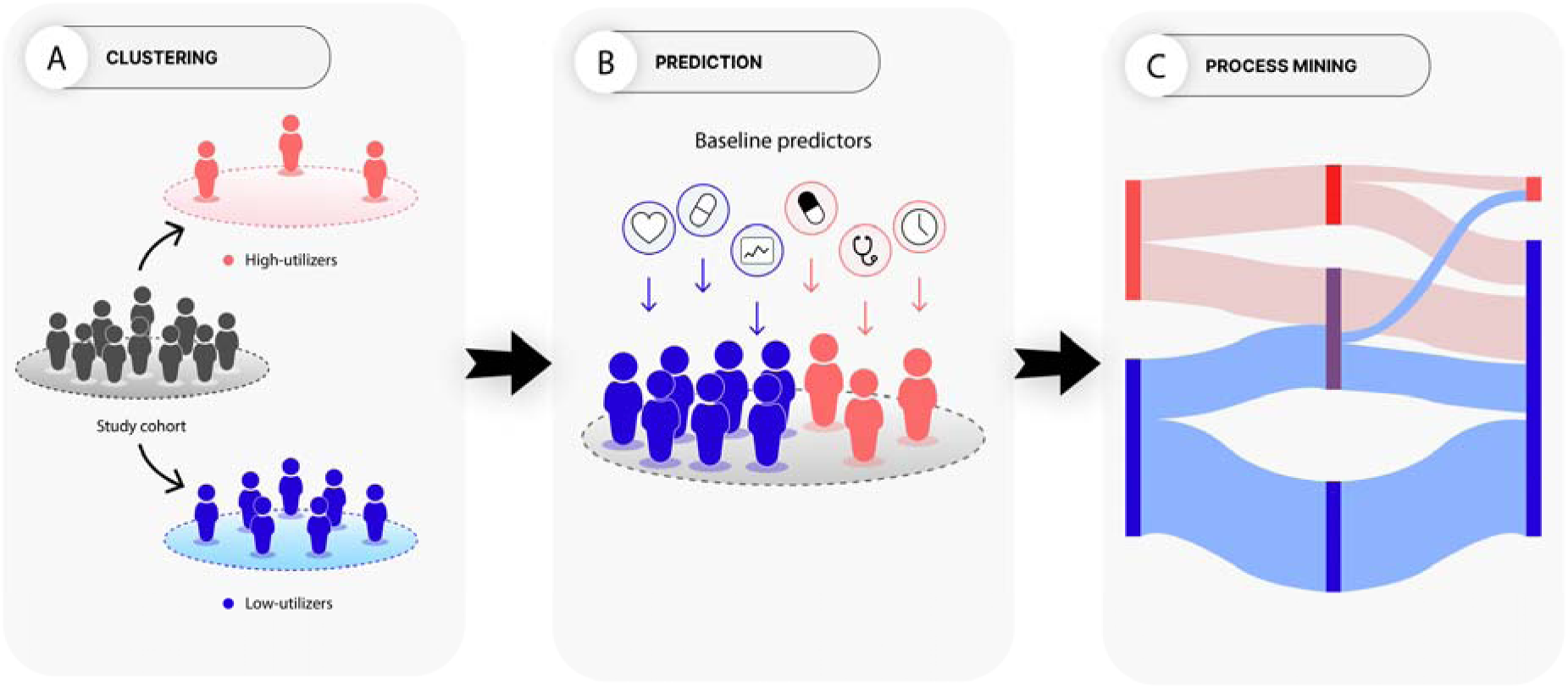
Study design and analytic workflow. Electronic health records were used to define the study cohort (N = 962) that was clustered in data-driven manner into two clusters based on long-term (years 2-5) healthcare utilization (A). Then, baseline demographic and clinical characteristics were entered as predictors in a logistic regression model to estimate the probability of cluster membership (B). Care trajectories were then mapped using process mining (C).

### Data and Preprocessing

The study cohort consisted of all patients diagnosed with RRMS (G35.A, ICD-10) between 1 January 2009 and 31 December 2018 reported to the clinician-curated MS quality registry (StellarQ; n =1005) [27]. Diagnosis was set by the treating neurologist. Patients were excluded if they were lacking all demographic data (n=7), lacked any encounter data (n=19) in the follow-up period, were under the age of 18 at diagnosis (n=16), or died during follow-up (n=1). Each patient was followed from one year before their diagnosis date (pre-diagnosis period) through five years post-diagnosis (follow-up period). Encounters beyond five years were censored to standardize follow-up time across patients.

For the study cohort, data were extracted from HUS administrative and clinical information systems (Apotti, Epic Systems Corporation, Wisconsin, USA, and Uranus, CGI, Montréal, Canada) for the period 1 January 2008 to 31 December 2023. For each patient, a chronological event log was assembled containing the patient identifier (de-identified for analysis), encounter start date (and end date for inpatient admissions), encounter type that was classified as (1) remote contact (RM) a non-physical contact such as telephone calls, secure messages, or e-visits; (2) outpatient visit (OP): a scheduled visits to neurology clinics; (3) emergency department visit (ED): visits to the emergency department; and (4) inpatient episode (IP): a hospital admissions to neurology wards. Only encounters within neurology services at HUS were included. Acute and unscheduled care utilization were defined as any ED or IP encounter, whereas RM and OP were defined as non-acute and scheduled care.

Baseline variables at diagnosis included age, sex, year of diagnosis, comorbidity burden (Charlson Comorbidity Index [CCI]) [28,29], disease severity (first recorded Expanded Disability Status Scale [EDSS]). Disease-modifying therapy (DMT) initiation post-diagnosis was recorded and categorized by efficacy (high, low-medium, other, no medication) for the first medication initiated (Table S1). Healthcare utilization was monetized from the payer’s perspective using the 2024 HUS tariff list (Table S2). All costs are expressed in 2024 euros (€) without discounting. These costs include service encounter costs and notably exclude pharmaceuticals. Costs were assigned to each encounter type recorded in the event logs. Each patient’s total cost over the study period was calculated by multiplying the number of encounters in each category by its respective unit cost. Inpatient admission costs were calculated as cost per day multiplied by length of stay.

### Data Analysis

#### Clustering Analysis

To identify distinct long-term healthcare utilization patterns, we vectorized each patient’s annual encounter data for follow-up years 2–5, excluding the initial diagnosis phase (years 0–1) to focus on established care trajectories. For each patient, a feature matrix was constructed, where each vector captured the year of follow-up, type of contact (inpatient, ED, outpatient, remote) and count of said contact. Variables were standardized (z-scores) to equalize scale across encounter types. K-means clustering was then applied to the standardized feature matrix.

The number of clusters (k) was determined using average silhouette width and total within-cluster sum of squares (elbow method). To assess clustering robustness, we performed a bootstrap stability analysis by repeatedly re-sampling 80% of the cohort (50 iterations) and re-running k-means clustering. Cluster agreement between resampled and original assignments was quantified using the adjusted Rand index (ARI). Mean and median ARI values were used to summarize stability. Clusters were labelled as high-utilization and low-utilization based on annual encounter volumes across all care settings.

#### Statistical Analysis

Continuous variables are presented as mean (standard deviation [SD]) and median [interquartile range (IQR)]. Categorical variables are reported as frequencies and percentages (n (%)). Univariate comparisons between clusters were performed using the Wilcoxon rank-sum test (Mann-Whitney U test) for continuous and ordinal variables, and Pearson’s chi-squared test for categorical variables. All p-values are two-sided, and a p-value < 0.05 was considered statistically significant.

Multivariable logistic regression was used to identify predictors of high-utilization cluster membership. The dependent variable was binary (high vs. low utilization, as determined by clustering). Predictor variables included baseline characteristics (sex, age at diagnosis [modeled as a restricted cubic spline with knots at 35 and 50 years], EDSS, CCI, diagnosis period, and efficacy category of first initiated medication), early-care utilization metrics from years 0–1: early scheduled contacts (sum of RM and OP encounters), early unscheduled contacts (sum of ED and IP encounters), and binary variables for early documented relapses (1 relapse, and ≥2 relapses). EDSS missingness was modeled as a separate binary indicator.

Odds ratios (ORs) with 95% confidence intervals (CIs) and p-values are reported. Model discrimination was assessed using both the area under the receiver operating characteristic curve (ROC AUC) and the area under the precision–recall curve (PR AUC) to account for moderate class imbalance (14.1% high utilizers). Multicollinearity among predictors was assessed using the generalized variance inflation factor (GVIF).

#### Process Mining

PM was used to visualize and quantify the longitudinal care trajectories that underlie the aggregate utilization counts used for clustering. This approach provided a complementary, sequence-based view of care patterns for the full cohort and stratified by cluster. All PM analyses were based on the event log constructed previously, using the de-identified patient identifier as the case ID, the encounter start date as the timestamp, and the four-category encounter type (remote, outpatient, ED, inpatient) as the activity.

First, to characterize the complexity and heterogeneity of care trajectories, we computed trace-level metrics including: (i) the total number of unique trajectories (traces); (ii) the trajectory coverage, defined as the cumulative proportion of patients represented by the 10 most common traces, and (iii) the average trajectory length, calculated as the mean number of events (encounters) per patient. These were calculated only for the post-diagnosis 5-year follow-up period, to focus the analysis on the period after a formal care plan is established, thereby better reflecting adherence to, or deviations from, intended care pathways.

Second, to visualize high-level care trajectories over time, each patient’s chronological sequence of encounters was transformed into a single “dominant care state” for each year of the study period (pre-diagnosis year 1 through post-diagnosis year 5). The dominant state was assigned based on a predefined hierarchy reflecting clinical acuity and resource intensity: inpatient > ED > high-intensity outpatient (≥3 contacts) > outpatient > remote > no contact. The time window of one year and the ≥3 outpatient contacts threshold was chosen based on clinical insight from two authors who work as neurologists based on managerial relevance and to distinguish low-frequency routine follow-up from more intensive scheduled care. We then constructed a Sankey diagram to depict the flow and volume of patients transitioning between these dominant states from the pre-diagnosis year through the five years post-diagnosis.

#### Ethical considerations

The study makes use of observational registry data. Under Finnish legislation (Finlex 552/2019), retrospective registry studies do not require informed consent. Approval for this study was obtained from HUS (permit number HUS 3411/2023).

## Results

### Study Population

A total of 962 patients with relapsing–remitting multiple sclerosis (RRMS) were included, contributing 5,772 person-years of follow-up. Figure 2 shows the distribution of incident RRMS cases by diagnosis year, stratified by cluster (left panel), and the total volume of healthcare contacts during the first post-diagnosis year (right panel). Clustering remained consistent across diagnosis years, and outpatient and remote encounters predominated throughout the observation period. Baseline characteristics are presented in Table 1. The mean age at diagnosis was 35.7 years (SD 9.8), and 71.7% (n = 690) were female, consistent with the known female predominance in MS. The median Charlson Comorbidity Index (CCI) at baseline was 0 with a mean of 0.1 (SD 0.5), and 49.7% (n = 479) experienced at least one documented clinical relapse during follow-up.

**Figure 2.**
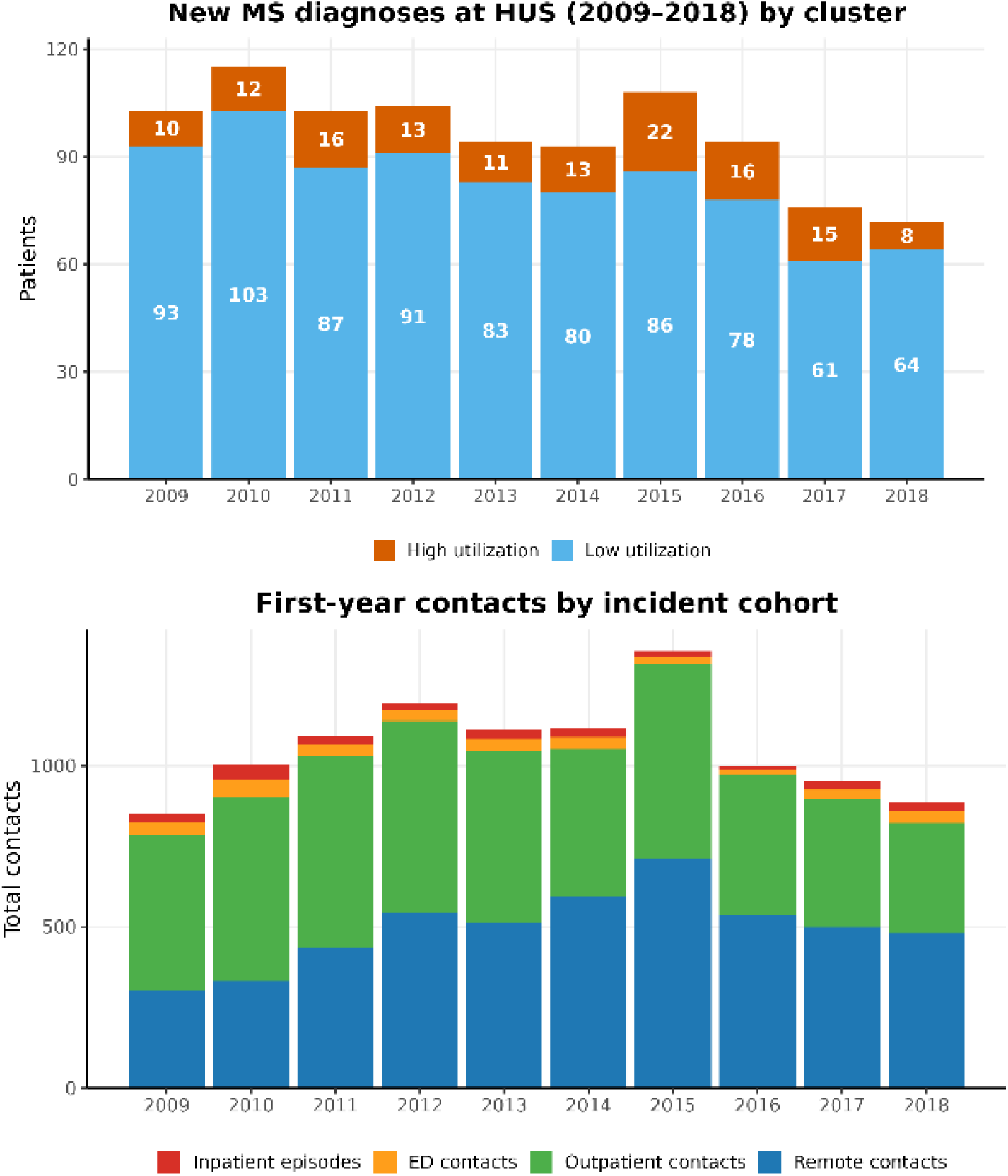
Annual number of new RRMS diagnoses at Helsinki University Hospital from 2009–2018, stratified by cluster (upper) and first-year (post-dx) healthcare contacts for the incident cohort, by care setting (lower).

**Table 1.** Baseline characteristics and annual healthcare utilization of the study cohort (n = 962) stratified by cluster. Values are presented as n (%), mean (SD), or median [interquartile range], as appropriate. Healthcare utilization variables represent annual contact frequency or cost. Encounter types: remote contacts (e.g., telephone, e-visits, secure messages); outpatient visits; emergency department visits; inpatient episodes. Costs are in 2024 euros (€), from the payer’s perspective, based on the Helsinki University Hospital 2024 tariff list. Median annual costs were approximately threefold higher in the high-utilization cluster than in the low-utilization cluster.

| Variable | Low utilization<br>(n=826) | High utilization<br>(n=136) | P-value | Full Cohort<br>(n=962) |
| --- | --- | --- | --- | --- |
| **Sex, n (%)** |  |  | 0.911 |  |
| Female | 593 (71.8) | 97 (71.3) |  | 690 (71.7) |
| Male | 233 (28.2) | 39 (28.7) |  | 272 (28.3) |
| **Age at diagnosis, years** |  |  | 0.001 |  |
| Mean (sd) | 36.2 (9.9) | 33.1 (8.7) |  | 35.7 (9.8) |
| **Charlson comorbidity index** |  |  | 0.037 |  |
| Median [IQR] | 0 [0.0 – 0.0] | 0 [0.0 – 0.0] |  | 0 [0.0 – 0.0] |
| Mean (sd) | 0.13 (0.50) | 0.21 (0.57) |  | 0.1 (0.5) |
| **Documented relapse** |  |  |  |  |
| Patients with ≥ 1 relapses (%) | 376 (45.5) | 102 (75.0) | < 0.001 | 478 (49.7) |
| Relapses per patient, median [IQR] | 1 [1–2] | 3 [1–4] | < 0.001 | 1 [1–2] |
| Patients with 1 early (years 0-1) relapse (%) | 199 (24.1) | 33 (24.3) | 0.965 | 232 (24.1) |
| Patients with ≥ 2 early (years 0-1) relapses (%) | 76 (9.2) | 51 (37.5) | < 0.001 | 127 (13.2) |
| **First recorded EDSS** |  |  | < 0.001 |  |
| Median [IQR] | 2.0 [1.0–2.5] | 2.5 [1.5–4.0] |  | 2.0 [1.0 – 2.0] |
| Missing (%) | 180 (21.8) | 15 (11.0) |  | 195 (20.3) |
| **Diagnosis period, n (%)** |  |  | 0.082 |  |
| 2009-2011 | 283 (34.3) | 38 (27.9) |  | 321 (32.4) |
| 2012-2014 | 254 (30.8) | 37 (27.2) |  | 291 (30.2) |
| 2015-2018 | 289 (35.0) | 61 (44.9) |  | 350 (36.4) |
| **Annual healthcare utilization (post-dx)** |  |  |  |  |
| Remote contacts, median [IQR] | 2.0 [1.0–4.8] | 7.0 [3.0–10.0] | < 0.001 | 3.0 [1.0 – 5.0] |
| Remote contacts, mean (sd) | 3.02 (3.2) | 7.42 (5.6) | < 0.001 | 3.6 (4.0) |
| Outpatient visits, median [IQR] | 1.0 [1.0–2.0] | 7.0 [3.0–13.0] | < 0.001 | 1.0 [1.0 – 3.0] |
| Outpatient visits, mean (sd) | 1.98 (2.3) | 8.26 (6.1) | < 0.001 | 2.9 (3.8) |
| Emergency department, median [IQR] | 0.0 [0.0–0.0] | 0.0 [0.0–0.0] | < 0.001 | 0.0 [0.0 – 0.0] |
| Emergency department, mean (sd) | 0.11 (0.6) | 0.49 (1.4) | < 0.001 | 0.2 (0.7) |
| No emergency department contacts (%) | 679 (82.2) | 72 (52.9) | < 0.001 | 751 (78.0%) |
| Inpatient episodes, median [IQR] | 0.0 [0.0–0.0] | 0.0 [0.0–0.0] | < 0.001 | 0.0 [0.0 – 0.0] |
| Inpatient episodes, mean (sd) | 0.06 (0.3) | 0.30 (0.7) | < 0.001 | 0.1 (0.4) |
| No inpatient episodes (%) | 626 (75.8) | 48 (35.3) | < 0.001 | 674 (70.0%) |
| **Annual cost (post-dx), € 2024** |  |  | < 0.001 |  |
| Median [IQR] | 629 [339–1209] | 2793 [1676–4249] |  | 774.0 [339 – 1578] |
| Mean (sd) | 1061 (1901) | 3808 (3992) |  | 1449.2 (2504) |
| **Total cost, € 2024** |  |  | < 0.001 |  |
| Median [IQR] | 6052 [3915-9634] | 18746 [15634-24738] |  | 7188 [4240-12040] |
| Mean (sd) | 7421 (6024) | 22527 (13180) |  | 9556 (9126) |

After diagnosis, patients had a median of 1.0 neurologist outpatient visits per year (IQR 1.0–3.0) and 3.0 remote contacts per year (IQR 1.0–5.0) annually, most commonly for routine follow-up, treatment monitoring, or administrative communication. Emergency department (ED) use was low, with an annual mean of 0.2 visits (SD 0.7; median 0), and most patients had no ED (78.0 %) contacts at all. Inpatient admissions were similarly infrequent, with a mean of 0.1 (SD 0.4; median 0) episodes per year and 70.0% of patients had no inpatient episodes during the follow-up period. Overall, outpatient encounters accounted for the vast majority of healthcare contacts in this cohort. These patterns highlight that RRMS management at HUS was largely ambulatory, punctuated by relatively infrequent acute-care episodes.

### Clustering Analysis

Two distinct long-term healthcare utilization clusters emerged from k-means clustering of encounter counts during follow-up years 2–5 (Figure 3). Cluster 1 (n = 136, 14.1%) was characterized by persistently high annual encounter volumes across all care settings, with median [IQR] annual outpatient and remote encounters of 7.0 [3.0–13.0] and 7.0 [3.0–10.0], respectively.

**Figure 3.**
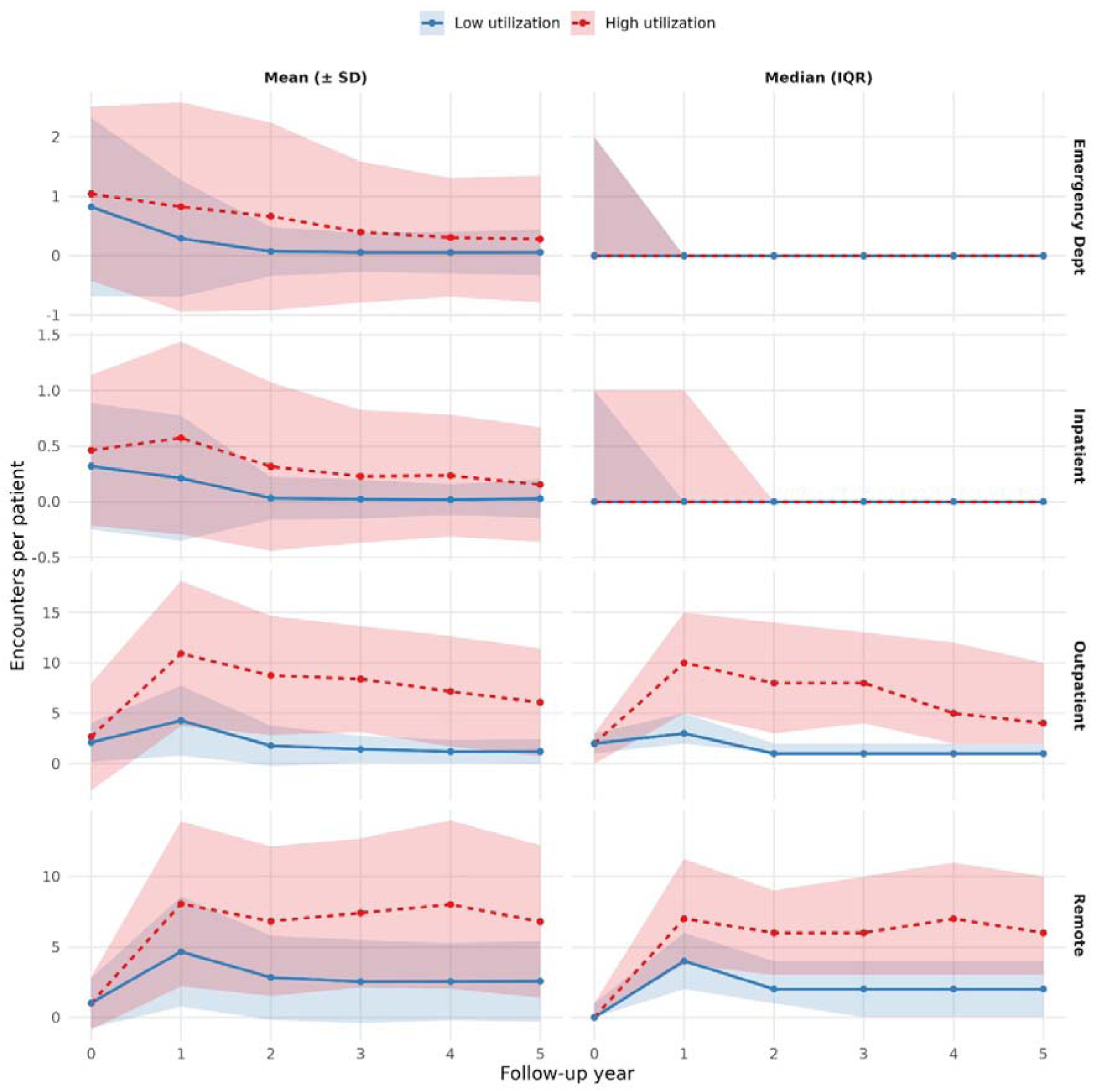
Annual healthcare utilization stratified by clusters. Clusters were defined using utilization data from years 2–5. Plots show per-patient mean (left) and median (right) annual counts of remote contacts, outpatient contacts, emergency department contacts, and inpatient episodes stratified by the two utilization clusters (low vs high).

Cluster 2 (n = 826, 85.9%) had markedly lower annual encounter volumes, with median 1.0 [1.0–2.0] outpatient encounters and 2.0 [1.0–4.8] remote encounters. While the total within-cluster sum of squares (WSS) showed a steady decline with increasing *k*, without a pronounced elbow, the average silhouette width peaked at 0.54 for *k* = 2, supporting it as a parsimonious solution (Figure S1). The two-cluster solution was robust to resampling (mean and median adjusted Rand index = 0.85), indicating consistent recovery of the high- and low-utilization clusters (Figure S2).

Total service encounter related costs over the study period differed substantially between the two clusters (Table 1). The median total cost per patient was €18,746 [IQR 15634–24738] in the high-utilization cluster compared with €6,052 [IQR 3915–9634] in the low-utilization cluster (p < 0.001, Wilcoxon rank-sum test). Mean (±SD) costs showed a similar pattern (€22,527 ± 13,180 vs €7421 ± 6,024), corresponding to an approximate threefold cost difference.

Outpatient and remote contacts accounted for the majority of total expenditures in both clusters, but high utilizers also incurred higher inpatient and ED costs (Figure 4). While inpatient episodes and ED contacts represented a smaller share of total contacts, they contributed disproportionately to overall cost among high utilizers.

**Figure 4.**
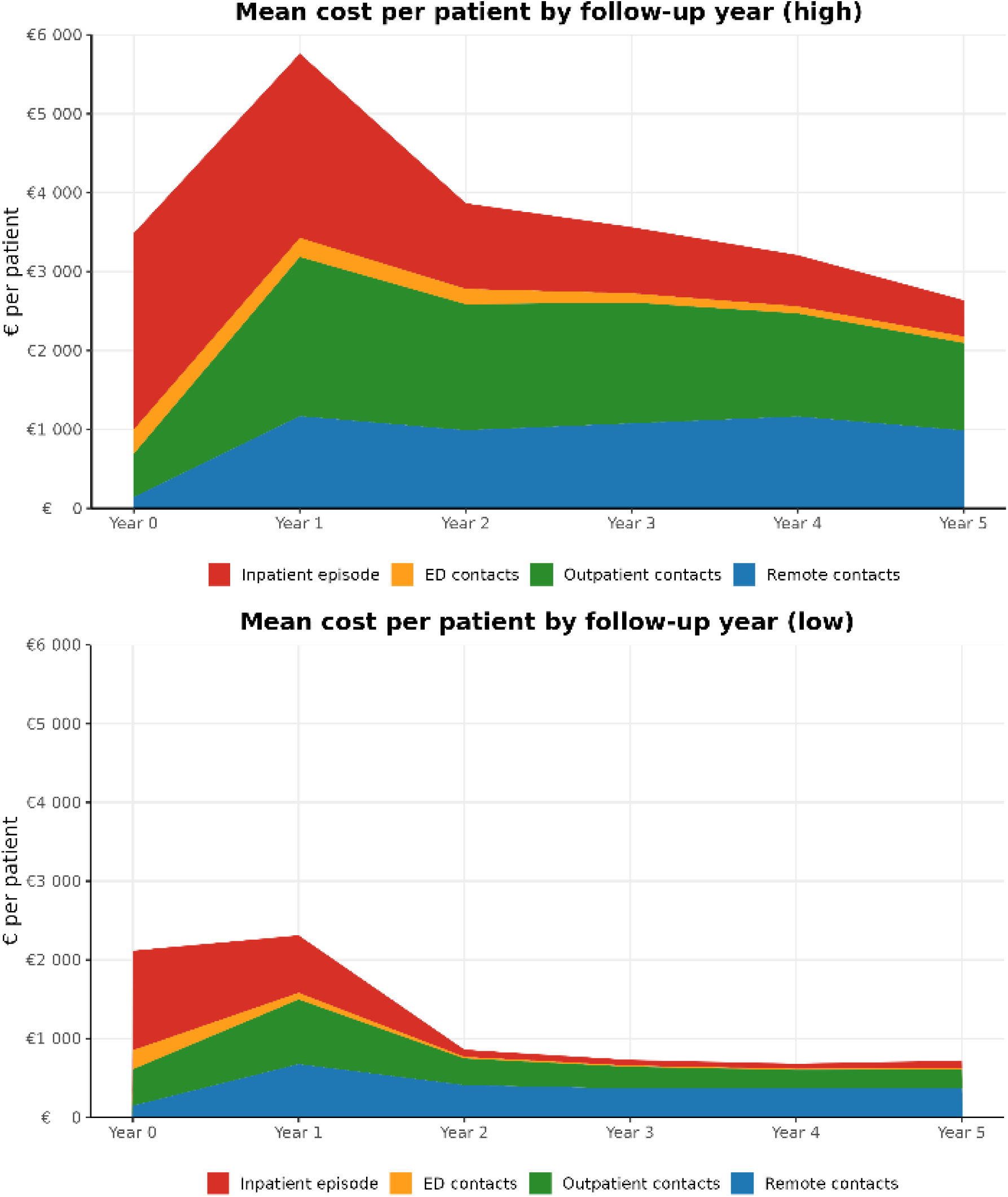
Stacked area plots showing the mean total direct healthcare cost per patient, in €2024, stratified by utilization cluster over the six-year study period. The high utilization cluster (upper; n=136) exhibits substantially higher overall costs, with inpatient (red) costs comprising a larger proportion compared to the low utilization cluster (lower; n=826). In both clusters, outpatient (green) and remote (blue) contacts account for the majority of expenditures.

### Logistic Regression Analysis

Multivariable logistic regression identified several baseline and early-care predictors for a patient to belong into the high-utilization cluster (Table 2). Experiencing two or more early relapses was the strongest predictor of high utilization (OR = 6.33, 95% CI 3.49–11.50, p < 0.001). Early scheduled contacts (remote and outpatient) in years 0–1 were strongly associated with later high utilization (OR = 1.07, 95% CI 1.04–1.10, p < 0.001), whereas early unscheduled contacts (ED and inpatient episodes) were not significant predictors (OR = 0.96, 95% CI 0.88–1.04, p = 0.320). Higher baseline EDSS scores were associated with increased odds of high utilization (OR = 1.50, 95% CI 1.27–1.78, p < 0.001), while missing EDSS data was not significantly associated with utilization (OR = 0.53, 95% CI 0.27–1.04, p = 0.060).

**Table 2.** Predictors of high-utilization cluster membership in multivariable logistic regression. Notes: OR = odds ratio; CI = confidence interval. Reference categories: sex = female, diagnosis period = 2009–2011, early relapses = 0, age modeled as a restricted cubic spline with knots at 35 and 50 years, first medication category = “high-efficacy DMT”. “Early scheduled contacts” = total remote and outpatient encounters in years 0–1. “Early unscheduled contacts” = total ED and inpatient encounters in years 0–1. EDSS missingness coded as separate category. Significant associations (p < 0.05) in bold.

| Variable | Estimate | Std. Error | OR (95 % CI) | p value |
| --- | --- | --- | --- | --- |
| **Early scheduled contacts** | **0.069** | **0.015** | **1.07 (1.04–1.10)** | **<0.001** |
| Early unscheduled contacts | -0.044 | 0.044 | 0.96 (0.88–1.04) | 0.320 |
| Male sex | 0.033 | 0.269 | 1.03 (0.60–1.74) | 0.903 |
| Age, linear term | -0.019 | 0.029 | 0.98 (0.93–1.04) | 0.513 |
| Age, spline knot (>35 y) | -0.058 | 0.053 | 0.94 (0.85–1.05) | 0.280 |
| Age, spline knot (>50 y) | -0.076 | 0.144 | 0.93 (0.65–1.33) | 0.599 |
| **EDSS score** | **0.404** | **0.086** | **1.50 (1.27–1.78)** | **<0.001** |
| EDSS missing | -0.632 | 0.336 | 0.53 (0.27–1.04) | 0.060 |
| Charlson comorbidity index | -0.076 | 0.213 | 0.93 (0.59–1.38) | 0.720 |
| **Medication category: Low-Medium** | **-2.411** | **0.347** | **0.09 (0.04–0.18)** | **<0.001** |
| **Medication category: No Medication** | **-3.062** | **0.702** | **0.05 (0.01–0.23)** | **<0.001** |
| **Medication category: Other** | **-1.441** | **0.674** | **0.24 (0.06–0.97)** | **0.033** |
| Diagnosis period: 2012–2014 | -0.128 | 0.313 | 0.88 (0.48–1.62) | 0.683 |
| Diagnosis period: 2015–2018 | 0.185 | 0.289 | 1.20 (0.68–2.13) | 0.521 |
| One early relapse | 0.212 | 0.301 | 1.24 (0.68–2.27) | 0.481 |
| **Two or more early relapses** | **1.845** | **0.304** | **6.33 (3.49–11.50)** | **<0.001** |

Compared to patients initiating high-efficacy DMTs, those starting on low-to-moderate efficacy therapies (OR = 0.09, 95% CI 0.04–0.18, p < 0.001), receiving no DMT (OR = 0.05, 95% CI 0.01–0.23, p < 0.001) or receiving “Other” medications (OR = 0.24, 95% CI 0.06–0.97, p = 0.033) all had substantially lower odds of high utilization, indicating confounding by indication. Other baseline characteristics, including sex, age at diagnosis, Charlson Comorbidity Index, and diagnosis period, were not significantly associated with high utilization.

The model’s discrimination was good, with ROC AUC = 0.89 (95% CI 0.86–0.92) and strong minority-class performance, with PR AUC = 0.64 (prevalence = 0.141), indicating over 4-fold enrichment over baseline (Figure S3). All generalized variance inflation factor (GVIF) scores were below 2 (< 3 for age variables), indicating a stable model with no multicollinearity issues. Predicted probabilities showed clear separation between clusters. However, there was notable overlap in the low predicted probability range (Figure S4).

### Process Mining Analyses

Overall, the full cohort exhibited extremely high process heterogeneity, with care trajectories in the high-utilization cluster being substantially longer and more varied (Table 3). The full cohort (n=962) contained 923 unique trajectories, meaning 95.9% of patients had a unique care trajectory. The 10 most common trajectories covered only 4.6% of all patients. Stratifying by cluster, the high-utilization group’s trajectories were, on average, 3.2 times longer (mean length 82.4 vs 25.9 events) and involved a wider variety of care services (mean unique activities 3.1 vs 2.4).

**Table 3.** Trace-level process mining metrics by utilization cluster.

| Metric | Low utilization (n=826) | High utilization (n=136) | Full Cohort (n=962) |
| --- | --- | --- | --- |
| Unique trajectories (traces) | 787 | 136 | 923 |
| Average count of unique activities, mean (sd) | 2.4 (0.7) | 3.1 (0.8) | 2.5 (0.8) |
| Average trajectory length (events), mean (sd) | 25.9 (14.9) | 82.4 (23.8) | 33.9 (25.7) |
| Coverage (top 10 traces), n (%) | 44 (5.3) | 10 (7.4) | 44 (4.6) |

To visualize how care trajectories evolved, we constructed Sankey diagrams mapping the flow of patients between yearly “dominant care states” from the pre-diagnosis year (Year 0) through five years post-diagnosis (Figure 5). These diagrams reveal two starkly different longitudinal patterns corresponding to the low- and high-utilization clusters.

**Figure 5.**
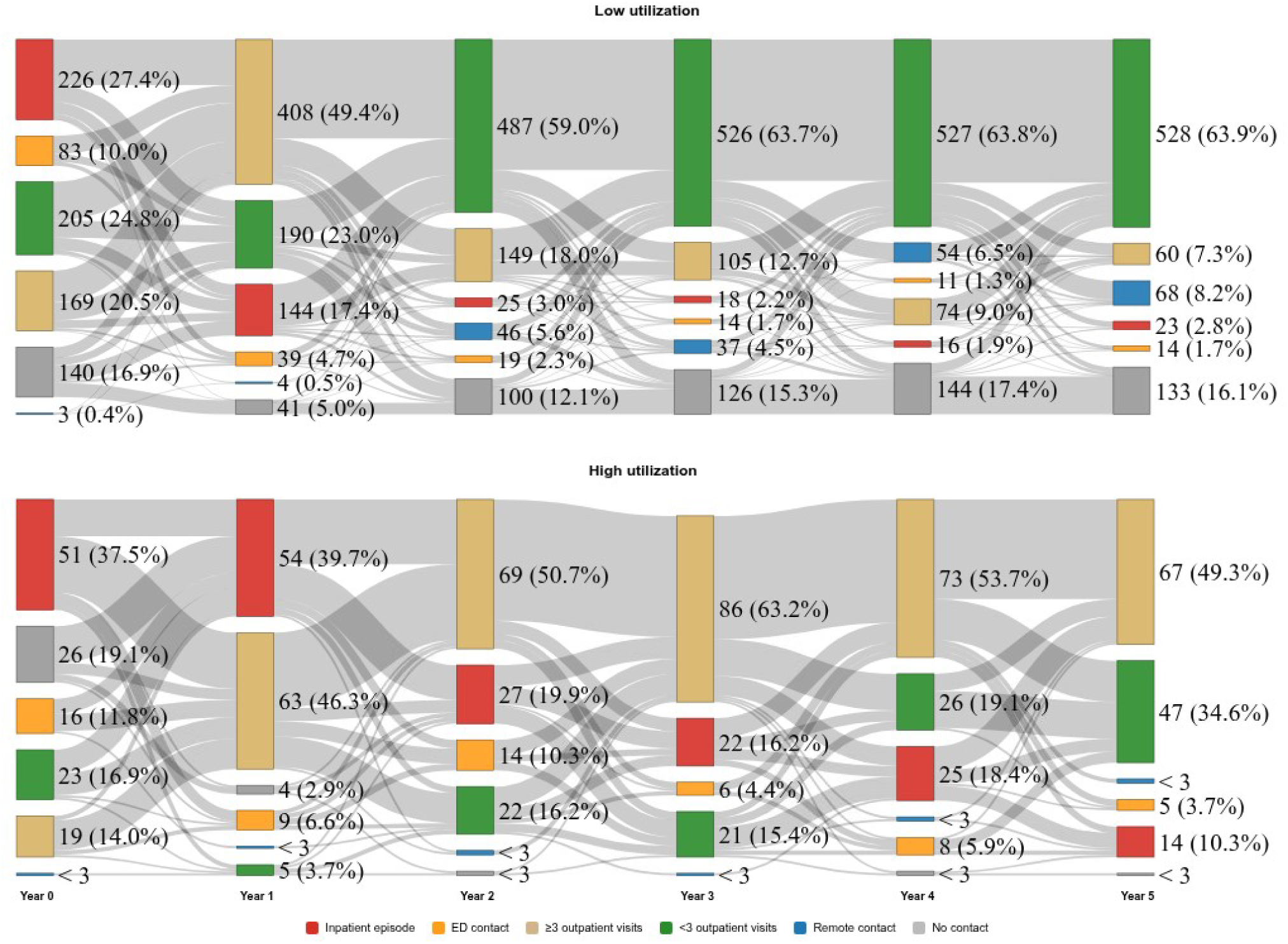
Patient flow between dominant yearly care states from one year before to five years after diagnosis in low utilization cluster (upper panel) and high utilization cluster (lower panel). Each patient was assigned a dominant care state for each study year according to the most resource-intensive encounter type recorded, using the following hierarchy: inpatient episode (red), emergency department contact (orange), ≥3 outpatient visits (tan), <3 outpatient visits (green) > remote contact only (blue) > no recorded contact (grey). Flows represent the number of patients transitioning between states in consecutive years; line width is proportional to the number of patients. Year 0 represents the year before diagnosis. Values where n < 3 are censored for GDPR compliance.

The low-utilization cluster’s trajectory (Figure 5) is defined by a rapid transition from high-acuity diagnostic care in years 0 and 1 to a stable, low-intensity state. In the pre-diagnosis year and first year after diagnosis, this group presented with a mix of states, including a large proportion of inpatient episodes (red) and high-intensity outpatient contacts (tan). However, following diagnosis, there is an immediate consolidation. By Year 2 post-diagnosis, the share of the inpatient episodes drops sharply, and the majority of the cohort is in the outpatient care state (green; 59.0%). In subsequent years, this pattern solidifies. By years 3-5 this stability is maintained, with the outpatient state becoming the clear dominant pathway for the cluster, ending with 63.9% of patients in this state by Year 5. Critically, the high-acuity inpatient episodes (red) and ED (orange) states, which were common at diagnosis, become negligible, accounting for only 2.8% and 1.7% of the cluster in Year 5, respectively. Similarly, the proportion of patients with no contacts (grey) in each year increases with time growing from 12.1% in Year 2 to 16.1% in Year 5.

In contrast, the high utilization cluster’s trajectory is characterized by persistent high-acuity and high-intensity care states. This clusters pre-diagnosis year was already dominated by inpatient episodes (red, 37.5%). Unlike the low-utilization group, these high-acuity states do not resolve after diagnosis. The inpatient state remains a major state for this cohort, accounting for 39.7% in Year 1, 19.9% in Year 2, and remaining a substantial pathway through all five years. The most prominent state for this cluster is the high-intensity outpatient state (tan). This state grows from 46.3% in Year 1 to become the dominant state for the majority of the follow-up period (e.g., 63.2% in Year 3). The stable outpatient state, which defined the low-utilization group, remains a minority pathway for this cluster. It is also noteworthy that the “no contact” state almost disappears in the high-utilization cluster after diagnosis.

In summary, the Sankey diagrams visually confirm the cluster identities: the low-utilization group is effectively stabilized into a low-intensity, routine-care pathway, while the high-utilization group remains “stuck” in a persistent, high-cost cycle of inpatient and intensive outpatient care. Overall, process mining revealed that high-utilization patients maintained high-intensity care states for longer, while low-utilization patients transitioned rapidly to low-intensity or no contact states. These patterns complemented the clustering results and highlighted distinct longitudinal care trajectories.

## Discussion

This study set out to generate managerially actionable insight into the heterogeneity and predictability of long-term care trajectories in relapsing–remitting multiple sclerosis (RRMS). Specifically, we asked how RRMS patients can be segmented into distinct utilization patterns over several years, and which baseline and early-care factors predict membership in a high-utilization group. To address these questions, we analyzed a comprehensive, six-year specialist-care cohort using longitudinal utilization data combined with regression-based risk modeling and process mining to reconstruct and compare multi-year care trajectories. We identified two distinct long-term healthcare utilization clusters: a high-utilization cluster (14.1%) with consistently greater encounter volumes across all care settings, and a low-utilization cluster (85.9%) with substantially lower resource use. Two or more early relapses, baseline EDSS, and early outpatient and remote contacts emerged as a strong predictor of later high utilization, while early inpatient and ED care were not predictive. Process mining (PM) revealed that high utilizers followed longer, more varied, and acute-oriented care pathways, with more frequent transitions into inpatient and emergency care states. Tariff-based costing demonstrated fourfold higher annual (€3,808 vs €1,060) and threefold higher cumulative (€22,527 vs €7,421) service encounter related costs over the six-year study period for the high utilization cluster underscoring the economic implications of early trajectory identification.

Unsurprisingly, higher disease burden (higher EDSS) was associated with higher utilization [30], however comorbidity (higher CCI) was not. The association between high-efficacy DMTs and high utilization likely represents confounding by indication: patients with highly active disease are appropriately selected for aggressive therapy. Two or more early relapses within the care trajectory emerged as the variable most strongly associated with high utilization. The strong association between early scheduled care activity and sustained high utilization suggests a sticky trajectory and contrasts with patterns reported previously, where early unscheduled care often predicts future costs [31]. In MS, high early scheduled contact volumes likely reflect more intensive monitoring following active disease onset or complex treatment initiation. The finding that early unscheduled care (ED/Inpatient) was not predictive of later high utilization is encouraging; it suggests that acute episodes in the early diagnostic phase are not necessarily indicative of a long-term unstable trajectory, provided they are effectively managed.

Our findings extend prior MS health services research by combining longitudinal clustering with PM. Previous studies have proposed three to five clusters of care utilization [19,20,32]. While our two-cluster solution is less granular, it maximizes managerial relevance by allowing for easy to implement operational utility. By consolidating these into a binary framework (stable vs. volatile), we provide a clearer signal for capacity planning. Our “high-utilization” cluster mirrors the “high-consumption” groups identified by Roux et al., confirming that a small subset of patients drives a disproportionate share of resources and costs. This finding is known to hold across diagnoses [33]. However, our PM analysis adds new depth by visualizing exactly how these costs accrue: through a failure to transition out of high-intensity “dominant states” in the years following diagnosis.

Our results suggest four concrete levers for managing long-term MS care at the organizational level. First, simple early indicators (≥2 relapses in years 0–1 and a high volume of early scheduled contacts) can be used as a pragmatic segmentation rule to flag the patients likely to follow a highlZlutilization trajectory. This rule can be embedded in EHR dashboards to support proactive identification during the first postlZldiagnosis year. Second, the ability to predict long-term high utilization using data from before and just after diagnosis for a life-long chronic disease presents an opportunity for targeted interventions. For high-risk patients, resources such as nurse coordinators, rapid-access clinics, or closer neurologist follow-up could help prevent the “drift” into high-cost trajectories [34,35]. The cost gradient between clusters provides an order-of-magnitude estimate of the budget envelope available for such targeted interventions. Third, for the ∼85% of patients in the low-utilization cluster, the rapid stabilization of their trajectories supports the safety of streamlined, remote-hybrid follow-up models [25]. This aligns with value-based healthcare principles: focusing resources on high-need patients while avoiding over-servicing those with stable disease courses [22]. Fourth, the process mined visualizations of care pathways offer a shared language for clinicians and administrators [36–38]. Sankey diagrams clearly illustrate the “funnel” effect of successful care, where patients move from complex diagnostic states to stable maintenance and highlight the minority who remain “stuck” in high-intensity loops. This “big picture” view is essential for justifying investments in resources such as nurse coordinators or rapid-access clinics designed to stabilize these trajectories.

The strengths of this study include the comprehensive capture of all neurological care within a single provider organization, minimizing loss to follow-up. Combining k-means clustering with PM provided a multi-dimensional view of care trajectories, validated by actual tariff-based costing. However, limitations exist. First, this was a single-center study in Finland, and local care delivery models may limit generalizability. Second, as with any EHR study, missing variables (particularly EDSS) may bias results. Third, while k-means proved robust, it assumes spherical clusters; future work could explore density-based clustering and group-based trajectory modeling approaches to detect more irregular trajectory shapes or improve on the division between the two clusters presented here. Finally, identified predictors represent markers of risk rather than causal drivers.

Future work should focus on externally validating these findings in other healthcare systems. Incorporating patient-reported outcomes (PROs) alongside utilization data could provide richer insights into the patient-level drivers of high resource use. Additionally, prospective testing of targeted interventions for patients flagged as high risk could determine whether these strategies can reduce utilization while maintaining care quality.

MS care process data provides a readily available and early signal of future service utilization. We demonstrate that the majority of RRMS patients rapidly stabilize into a predictable, low-cost trajectory, while a distinct minority generates sustained, high-intensity demand. Employing these data-driven phenotypes to tailor care pathways offers a concrete strategy for managing the operational complexity of chronic disease.

## Declarations

## Acknowledgements

The authors acknowledge StellarQ Ltd for continuous support for MS research in Finland. The authors also gratefully acknowledge the collaboration and support of CGI, Valtteri Lipsanen, Arto Ihantoja, and Miikka Kiiski, whose engagement in the process mining activities contributed valuable insights to this study.

## Data availability statement

Due to general data protection regulation (GDPR), individual identifiable patient data of the study cannot be shared.

## Supplementary material

**Table S1.**
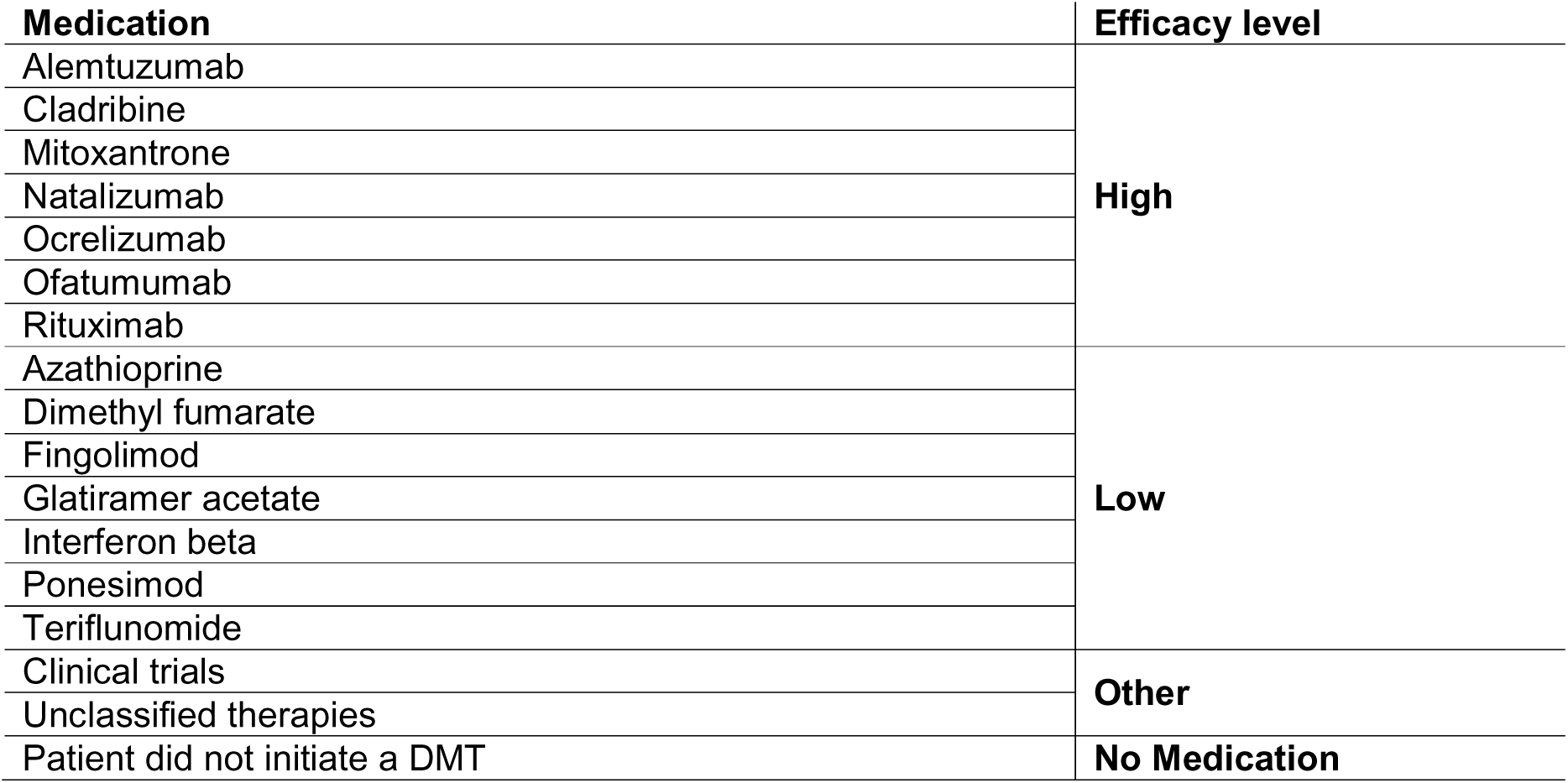
Classification of first Disease-Modifying Therapy (DMT) initiated post-diagnosis by efficacy.

**Table S2.** Unit costs (€LJ2024) applied to monetize healthcare utilization. Tariffs are drawn from the 2024 price list of Helsinki University Hospital and represent the payer’s perspective.

| Contact type | Unit cost |
| --- | --- |
| Inpatient episode | 802€ / day |
| ED contact (ED) | 296€ |
| First visit (Outpatient) | 296€ |
| Additional visit (Outpatient) | 175€ |
| Care visit (Outpatient) | 175€ |
| Serial visit (Outpatient) | 175€ |
| Call (Remote) | 145€ |
| Care call (Remote) | 145€ |
| Care mail (Remote) | 145€ |
| eDoctors visit (Remote) | 242€ |
| eSpecialist (Remote) | 175€ |

**Figure S1.**
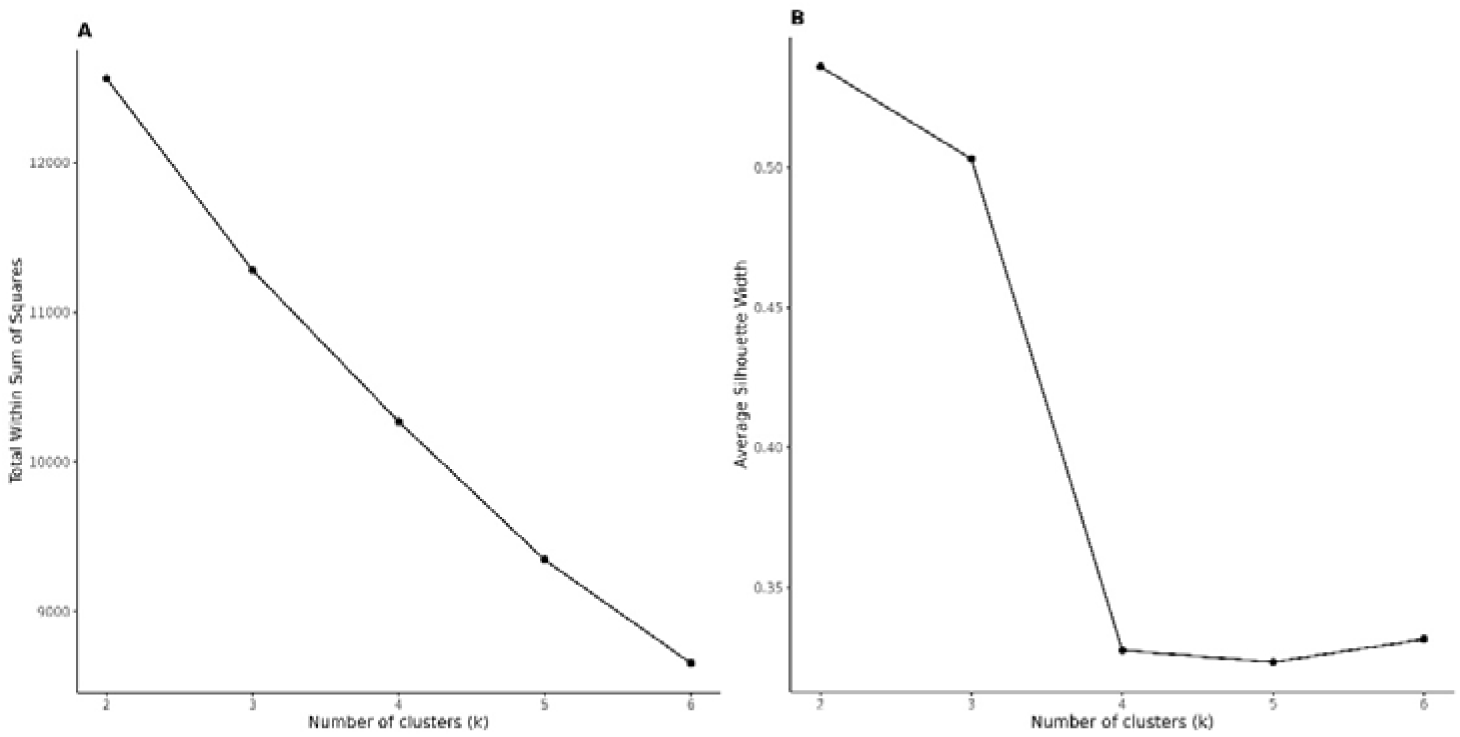
Cluster number selection using elbow and silhouette methods. Left panel: Elbow plot showing total within-cluster sum of squares (WSS) for k = 1–6; the inflection point occurs at k = 2–3. Right panel: Average silhouette width for k = 2–6, with the highest silhouette (∼0.54) at k = 2, supporting a two-cluster solution.

**Figure S2.**
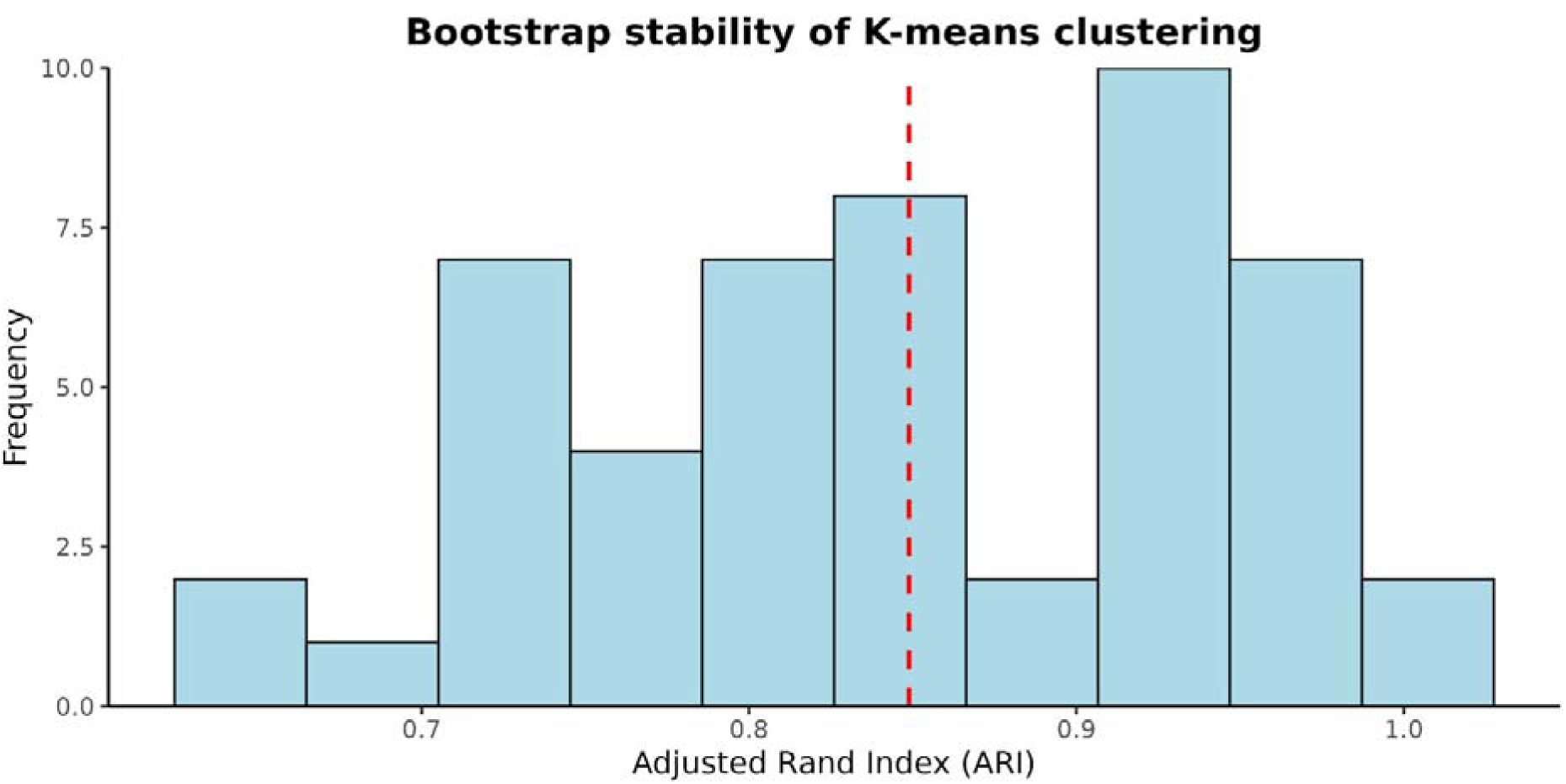
Bootstrap stability of k-means clustering. Distribution of adjusted Rand indices (ARI) from 50 bootstrap iterations. The mean ARI = 0.85 (red line) indicates strong reproducibility of the high- vs. low-utilization clusters.

**Figure S3.**
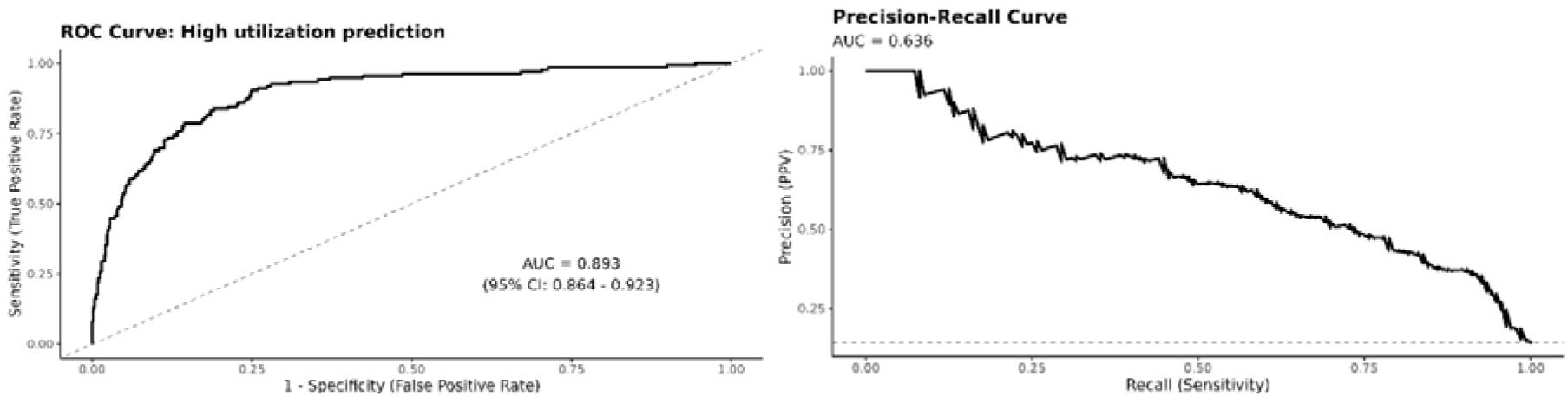
Area under the receiver operating characteristic (ROC AUC = 0.89) curve for logistic regression model (Left panel). Area under the precision–recall curve (PR AUC = 0.64) *for logistic regression model (right panel)*.

**Figure S4.**
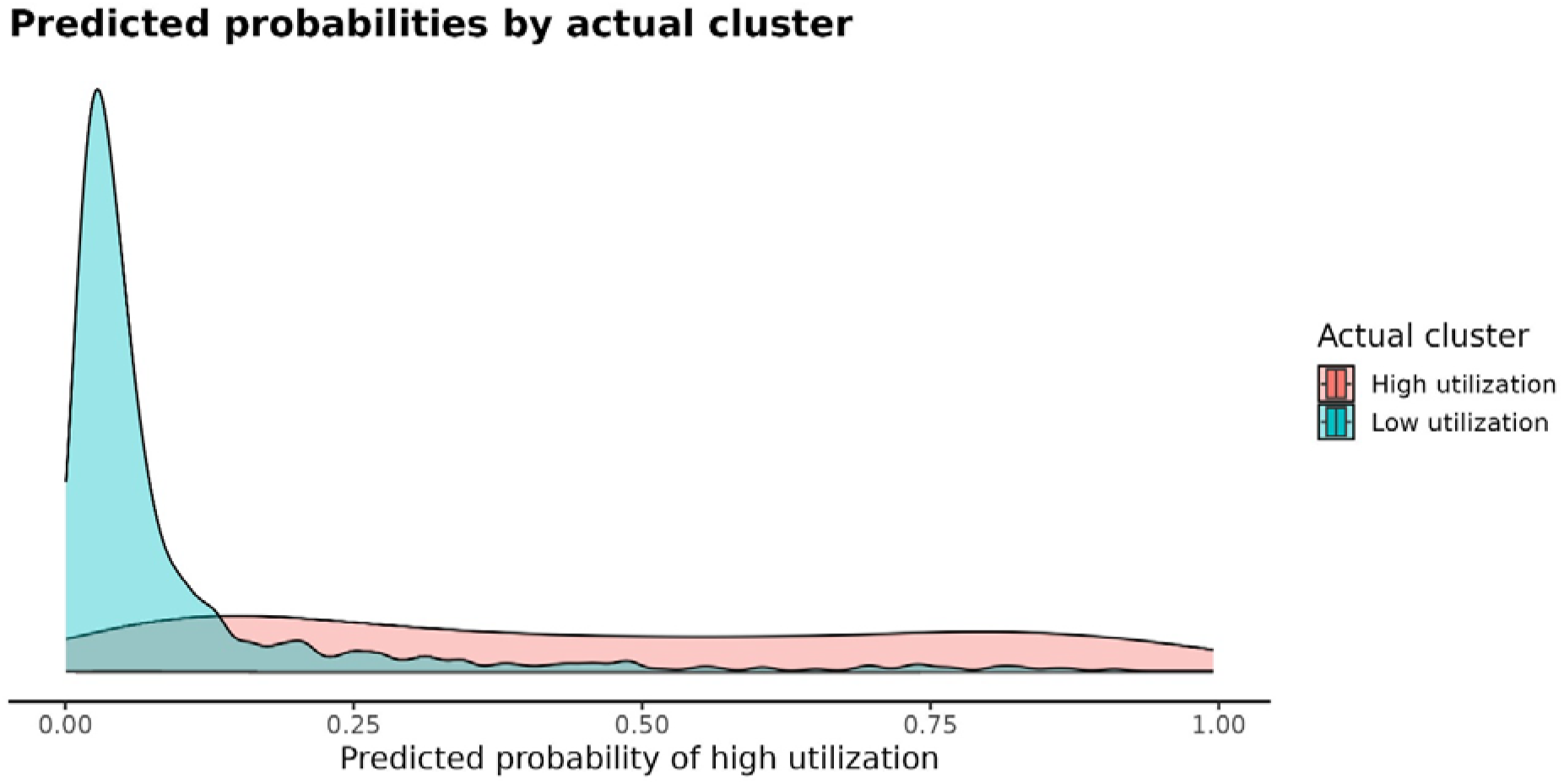
Distribution of predicted probabilities for high utilization by actual utilization cluster. The plot shows that individuals in the low utilization cluster (blue) have predicted probabilities concentrated near 0, while those in the high utilization cluster (red) have probabilities spread across higher values, indicating that the model generally distinguishes between the two clusters effectively, though with some overlap.

